# Vaccine effectiveness when combining the ChAdOx1 vaccine as the first dose with an mRNA COVID-19 vaccine as the second dose

**DOI:** 10.1101/2021.07.26.21261130

**Authors:** Mie Agermose Gram, Jens Nielsen, Astrid Blicher Schelde, Katrine Finderup Nielsen, Ida Rask Moustsen-Helms, Anne Katrine Bjørkholt Sørensen, Palle Valentiner-Branth, Hanne-Dorthe Emborg

**Affiliations:** Department of Infectious Disease Epidemiology and Prevention, Statens Serum Institut, Artillerivej 5, 2300 Copenhagen, Denmark; Department of Infectious Disease Preparedness, Data Integration and Analysis, Statens Serum Institut, Artillerivej 5, 2300 Copenhagen, Denmark

## Abstract

**Background:** The recommendations in several countries to stop using the ChAdOx1 vaccine has led to vaccine programs combining different vaccine types, which necessitates new knowledge on vaccine effectiveness (VE). In this study, we aimed to estimate the VE when combining the ChAdOx1 vaccine as the first dose and an mRNA vaccine as the second dose.

**Methods:** This nationwide population-based cohort study estimated VE against SARS-CoV-2 infection, all-cause and COVID-19 related hospitalization and death after receiving the ChAdOx1 vaccine as the first dose followed by an mRNA vaccine as the second dose. VE estimates were obtained using a Cox regression with calendar time as underlying time and adjusted for sex, age, comorbidity, heritage and hospital admission. Information on all individuals was extracted and linked from high-quality national registries.

**Results:** A total of 5,542,079 individuals were included in the analyses (97.6% of the total Danish population). A total of 144,360 were vaccinated with the ChAdOx1 vaccine as the first dose and of these 136,551 individuals received an mRNA vaccine as the second dose. A total of 1,691,464 person-years and 83,034 cases of SARS-CoV-2 infection were included. The VE against SARS-CoV-2 infection when combining the ChAdOx1 and an mRNA vaccine was 88% (95% confidence interval (CI): 83; 92) 14 days after the second dose and onwards. There were no COVID-19 related hospitalizations and deaths among the individuals vaccinated with the combination of the ChAdOx1 and an mRNA vaccine during the study period.

**Conclusion:** In conclusion, this study found a reduction in the risk of SARS-CoV-2 infection when combining the ChAdOx1 and an mRNA vaccine, compared with unvaccinated individuals. This is similar to the VE of two doses of an mRNA vaccine. Longer follow-up time is needed to confirm vaccine induced protection against severe events, such as COVID-19 related hospitalization and death.

## Introduction

The COVID-19 pandemic caused by the outbreak of the novel coronavirus SARS-CoV-2 in Wuhan, China, has caused major global public health concerns. As of 26^th^ of July 2021, more than 192 million cases and 4,1 million deaths have been reported worldwide (1). In Denmark, the COVID-19 vaccination program started on 27^th^ of December 2020, with the BNT162b2 mRNA vaccine from Pfizer/BioNTech, also called Comirnaty^®^. The mRNA-1273 vaccine (SpikeVax^®^ from Moderna) and the viral vector ChAdOx1 vaccine (Vaxzevria^®^ from AstraZeneca) were introduced in the vaccination program on 14^th^ of January and 9^th^ of February 2021, respectively. The ChAdOx1 vaccine was primarily given to front-line personnel in healthcare, elderly care and selected parts of the social sector who were at particular risk of infection with SARS-CoV-2 or who had been identified as performing critical functions in society (referred to as front-line personnel) (2). On 11^th^ of March 2021, it was decided to pause the use of the ChAdOx1 vaccine in the Danish COVID-19 vaccination program due to a possible link between the vaccine and very rare cases of unusual blood clots, bleeding and low blood platelet counts (3, 4). On April 14^th^ 2021, it was decided not to continue with the use of the ChAdOx1 vaccine in the Danish vaccination program, due to the present epidemiological situation. The Danish Health Authority announced on the 16^th^ of April 2021 that individuals who had only received the first dose of the ChAdOx1 vaccine should be offered a second dose of either the BNT162b2 mRNA or the mRNA-1273 vaccine (ChAdOx1/mRNA vaccine schedule) (4). Studies from the UK have reported VE estimates between 22% and 94% following the administration of one dose of ChAdOx1 (5-7). Due to changing recommendations regarding the use of the ChAdOx1 vaccine (4) and to avoid vaccine shortages, some countries are combining vaccine types. This creates a need for studies of VE for heterologous vaccination schedules (8). Immunological data on a heterologous vaccination schedule indicates that the combination of the ChAdOx1/mRNA vaccines is at least as immunogenic and protective as homologous BNT162b2 vaccination (9, 10). However, no previous studies have reported VE of a ChAdOx1/mRNA vaccine schedule. This study aimed to estimate VE against SARS-CoV-2 infection, all-cause and COVID-19 related hospitalization and death of 1) one dose of the ChAdOx1 vaccine and 2) the ChAdOx1/mRNA vaccine schedule, compared with unvaccinated individuals.

## Methods

### Study design and population

All residents in Denmark are registered in the Danish Civil Registration System and assigned a unique personal identification number (CPR number), which is used in all national registries, enabling accurate individual-level linkages between registries (11). In this nationwide retrospective population-based cohort study, individuals were included in the study population if they were residents in Denmark on 9^th^ of February 2021 or immigrated before the end of study on 23^rd^ of June 2021. Individuals who had received a COVID-19 vaccine or had a Reverse Transcription Polymerase Chain Reaction (RT-PCR) confirmed SARS-CoV-2 infection before the start of the study (2.4%) were excluded. The latter due to expected natural immunity from previousSARS-CoV-2 infection (12). The study participants were followed from 9^th^ of February 2021 and until a SARS-CoV-2 infection, receiving the first dose of another COVID-19 vaccine than the ChAdOx1 vaccine, emigration, death or end of follow-up (23^rd^ of June 2021), whichever came first. Information on immigration, emigration and vital status was retrieved from the Danish Civil Registration System (11). Information on the date of vaccination and type of vaccine was retrieved from the Danish Vaccination Registry, which includes data on all administered vaccines (13).

### Assessment of exposures

The general exposures of interest were: 1) a single dose of ChAdOx1 and 2) a second dose of either BNT162b2 mRNA or mRNA-1273 following the first dose of ChAdOx1. Unvaccinated individuals were used as reference. To examine the effect of one dose of ChAdOx1 on SARS-CoV-2 infection, time following vaccination was divided into 0-13 days (the run-in period), and 14 days and onward was divided into 7-day-intervals until receiving the second dose. The effect of one dose of ChAdOx1 on hospitalization and death was divided into 0-13 days (the run-in period) and 14 days and onwards days until receiving the second dose of an mRNA vaccine. The effect of the ChAdOx1/mRNA vaccine schedule on all outcomes was divided into 0-13 days and 14 days and onwards.

### Assessment of outcomes

The outcomes of interest were a SARS-CoV-2 infection, defined as a laboratory confirmed RT-PCR SARS-CoV-2 positive test, and all-cause or COVID-19 related hospitalization and death. Information on RT-PCR SARS-CoV-2 positive tests was retrieved from the Danish Microbiology Database (MiBa), which is a national database that automatically accumulates both positive and negative test results from all Danish departments of clinical microbiology (14). Information about rapid antigen tests was not included due to moderate sensitivity in asymptomatic patients compared with RT-PCR (22). A COVID-19 related hospitalization or death was defined as an admission within 14 days or death within 30 days after a positive SARS-CoV-2 test, respectively. Therefore, time at risk for the COVID-19 related outcomes were extended to 14 and 30 days after a confirmed SARS-CoV-2 infection, respectively. Hospital admissions and discharge dates were retrieved from the Danish National Patient Registry (15).

### Covariates

The incidence of SARS-CoV-2 varied considerably throughout the study period. To control for this variation through the study period, calendar time was used as underlying time. Further, age, sex, heritage and comorbidity were also included as covariates for the association between COVID-19 vaccination and SARS-CoV-2 infection, hospitalization and death. Finally, hospital admission was included as a covariate for the association between COVID-19 vaccination and SARS-CoV-2 infection and death. Information on date of birth and sex (male/female) was retrieved from the Danish Civil Registration System (11). The presence of comorbidity (yes/no) within the previous five years (data retrieved at start of study) was identified based on diagnoses coded according to the International Classification of Diseases, 10th revision (ICD-10). Diagnoses included one primary and optional secondary diagnosis for each patient contact and were retrieved from the Danish National Patient Registry (15). The ICD-10 codes included in comorbidity are provided in the supporting information (Table S1). Information on heritage (Danish/western/non-western/unknown) was retrieved from the Danish Civil Registration System (Table S2) (11). Only the heritage variable contained missing observations (0.2%); these were included as unknown.

## Statistical analysis

Characteristics of the included population were described using proportions. Crude VE estimates were obtained using a Poisson regression with overdispersion and time at risk as offset. Adjusted hazard ratios (HR) were obtained using a Cox regression model with calendar time as underlying time. The covariates sex, heritage and comorbidity were included as fixed covariates, being admitted to hospital as a time varying covariate, and age as a cubic spline. Time intervals after vaccination were included as a time varying covariate. Post estimation, VE estimates were calculated as 1 − HR · 100%. Data were analyzed using R version 4.0.5 (R Foundation for Statistical Computing, https://www.R-project.org/).

## Results

The study population included 5,542,079 individuals, among which 144,360 (2.6%) received the ChAdOx1 vaccine as the first dose. Of these, 88,050 (61%) and 48,501 (33.6%) received the BNT162b2 mRNA and the mRNA-1273 vaccine as the second dose, respectively. A total of 1,691,464 person-years and 83,034 SARS-CoV-2 infections were included. The study population had a median age of 45 and 46 years at the time of the first and second dose, respectively. There was an equal distribution of males and females. Comorbidity was present among 25.6% of the individuals and the majority had Danish heritage (84%) (Table 1).

**Table 1.** Characteristics of the study population.

|  |  | **N** | **%** |
| --- | --- | --- | --- |
| **Number of individuals included in the analyses** |  | 5,542,079 | 97.6 |
| **Median age (years) at first dose of the ChAdOx1 vaccine (IQR)** |  | 45 (33; 55) |  |
| **Median age (years) at second dose of an mRNA vaccine (IQR)** |  | 46 (34; 55) |  |
| **Sex** | Male | 2,801,776 | 50.6 |
|  | Female | 2,740,303 | 49.4 |
| **Comorbidity** | Yes | 1,416,332 | 25.6 |
|  | No | 4,125,747 | 74.4 |
| **Heritage** | Danish | 4,653,808 | 84.0 |
|  | Western | 369,255 | 6.7 |
|  | Non-western | 509,330 | 9.2 |
|  | Unknown | 9,686 | 0.2 |
| **Coverage of one dose of the ChAdOx1 vaccine** | Front-line personnel | 143,231 | 2.6 |
|  | Other | 1,129 | 0.02 |
| **Coverage of second dose of the BNT162b2 mRNA following a first dose ChAdOx1** |  | 88,050 | 1.6 |
| **Coverage of the second dose of the mRNA-1273 following a first dose ChAdOx1** |  | 48,501 | 0.9 |
| **Median follow-up days (min, IQR, max)** |  | 133 (1, 97, 135, 135) |  |
| **Median number of days to the first dose (IQR)** |  | 18 (11, 24) |  |
| **Median number of days from the first dose to the second dose (IQR)** |  | 82 (78, 85) |  |

The incidence of SARS-CoV-2 infection decreased from 27^th^ of December 2020 where the vaccination program was initiated in Denmark. Most likely, the decrease was a result of the combination of the newly started vaccination program and the lockdown from 16^th^ of December 2020 until 1^st^ of March 2021. The partial reopening in March 2021 resulted in a slight increase in SARS-CoV-2 infections until 1^st^ of June 2021, where approximately 35% and 20% of the population had received started or completed vaccination, respectively. As of 26th July 2021 74.3% of all SARS-CoV-2 RT-PCR positive test registered in 2021 had been sequenced. Based on the available sequencing data, the B.1.1.7 (alpha) variant was observed throughout the whole study and was dominant after mid-February. The B.1.617.2 (delta) variant first became prominent after the end of the present study (23^rd^ of June, 2021), and during the study period, only a small proportion of the delta variant was observed during the study period and (Fig 1).

**Fig 1.**
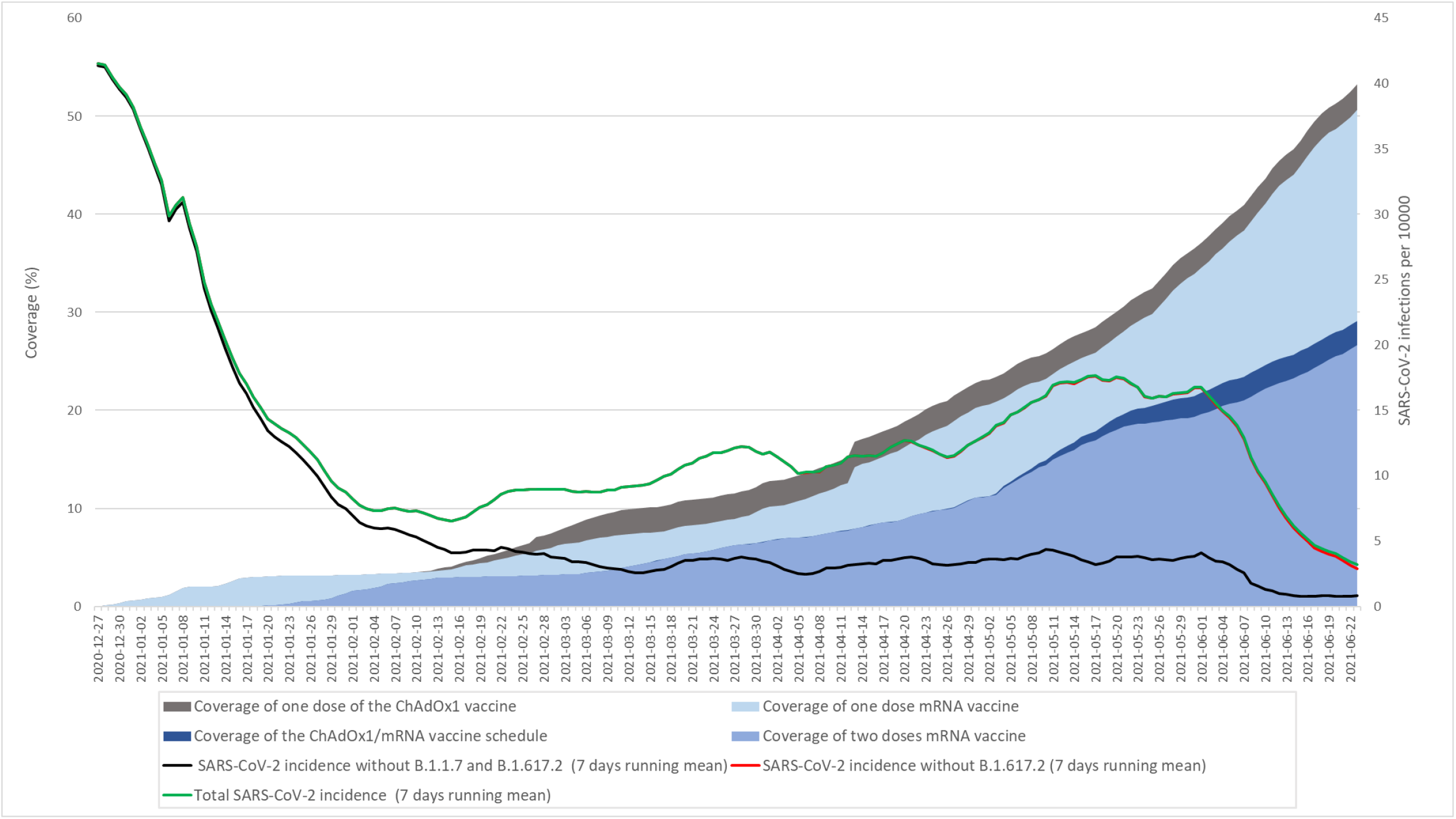
Percentage of population vaccinated and incidence of SARS-CoV-2 infection (7 days running mean)

Significant adjusted VE estimates against SARS-CoV-2 infection between 14-83 days after one dose of the ChAdOx1 vaccine were relatively stable with overlapping CIs. However, the VE estimates were not significant at 0-13 days, 42-48 days and from 84 days (Fig 2). The significant adjusted VE estimates ranged from 29% (95% CI: 12; 43) to 44% (95% CI: 32; 55) after one dose of the ChAdOx1 vaccine. For the ChAdOx1/mRNA vaccine schedule, significant adjusted VE estimates of 66% (95% CI: 59; 72) and 88% (95% CI: 83; 92) were observed at 0-13 days and from 14 days and onwards after the second dose, respectively (Table 2).

**Table 2.** Unadjusted and adjusted VE estimates against RT-PCR SARS-CoV-2 infection of one dose of the ChAdOx1 vaccine and the ChAdOx1/mRNA vaccine schedule.

| Vaccine type | Time interval (days) | No. of events | Person-years | Incidence rate | Unadjusted |  | Adjusted* |  |
| --- | --- | --- | --- | --- | --- | --- | --- | --- |
|  |  |  |  |  | VE, % | 95% CI | VE, % | 95% CI |
| Unvaccinated |  | 81,755 | 1,645,793 | 0,0497 | Reference |  | Reference |  |
| One dose of the ChAdOx1 vaccine | 0-13 | 197 | 5,528 | 0,0356 | 28 | 8; 44 | 0 | -15; 14 |
|  | 14-20 | 69 | 2,759 | 0,0250 | 50 | 23; 67 | 39 | 23; 52 |
|  | 21-27 | 66 | 2,758 | 0,0239 | 52 | 26; 69 | 44 | 29; 56 |
|  | 28-34 | 85 | 2,744 | 0,0310 | 38 | 8; 58 | 29 | 12; 43 |
|  | 35-41 | 81 | 2,741 | 0,0296 | 41 | 12; 60 | 35 | 19; 48 |
|  | 42-48 | 106 | 2,737 | 0,0387 | 22 | -10; 45 | 17 | -1; 31 |
|  | 49-55 | 96 | 2,733 | 0,0351 | 29 | -1; 51 | 29 | 3; 42 |
|  | 56-62 | 98 | 2,728 | 0,0359 | 28 | -3; 49 | 36 | 21; 47 |
|  | 63-69 | 99 | 2,660 | 0,0372 | 25 | -7; 47 | 41 | 28; 51 |
|  | 70-76 | 91 | 2,429 | 0,0375 | 25 | -9; 48 | 44 | 32; 55 |
|  | 77-83 | 83 | 1,723 | 0,0482 | 3 | -43; 34 | 31 | 14; 44 |
|  | 84-90 | 46 | 615 | 0,0748 | -51 | -154; 11 | -3 | -37; 23 |
|  | 91-97 | 12 | 237 | 0,0506 | -2 | -183; 63 | 23 | -36; 56 |
|  | 98-104 | 5 | 146 | 0,0342 | 31 | -235; 86 | 29 | -71; 70 |
|  | ≥105 | 7 | 161 | 0,0436 | 12 | -234; 77 | -47 | -208; 30 |
| The ChAdOx1/mRNA vaccine schedule | 0-13 | 109 | 5,197 | 0,0210 | 58 | 41; 70 | 66 | 59; 72 |
|  | ≥14 | 29 | 7,775 | 0,0037 | 92 | 86; 96 | 88 | 83; 92 |

\*Adjusted for age, sex, heritage, hospital admission and comorbidity

**Fig 2.**
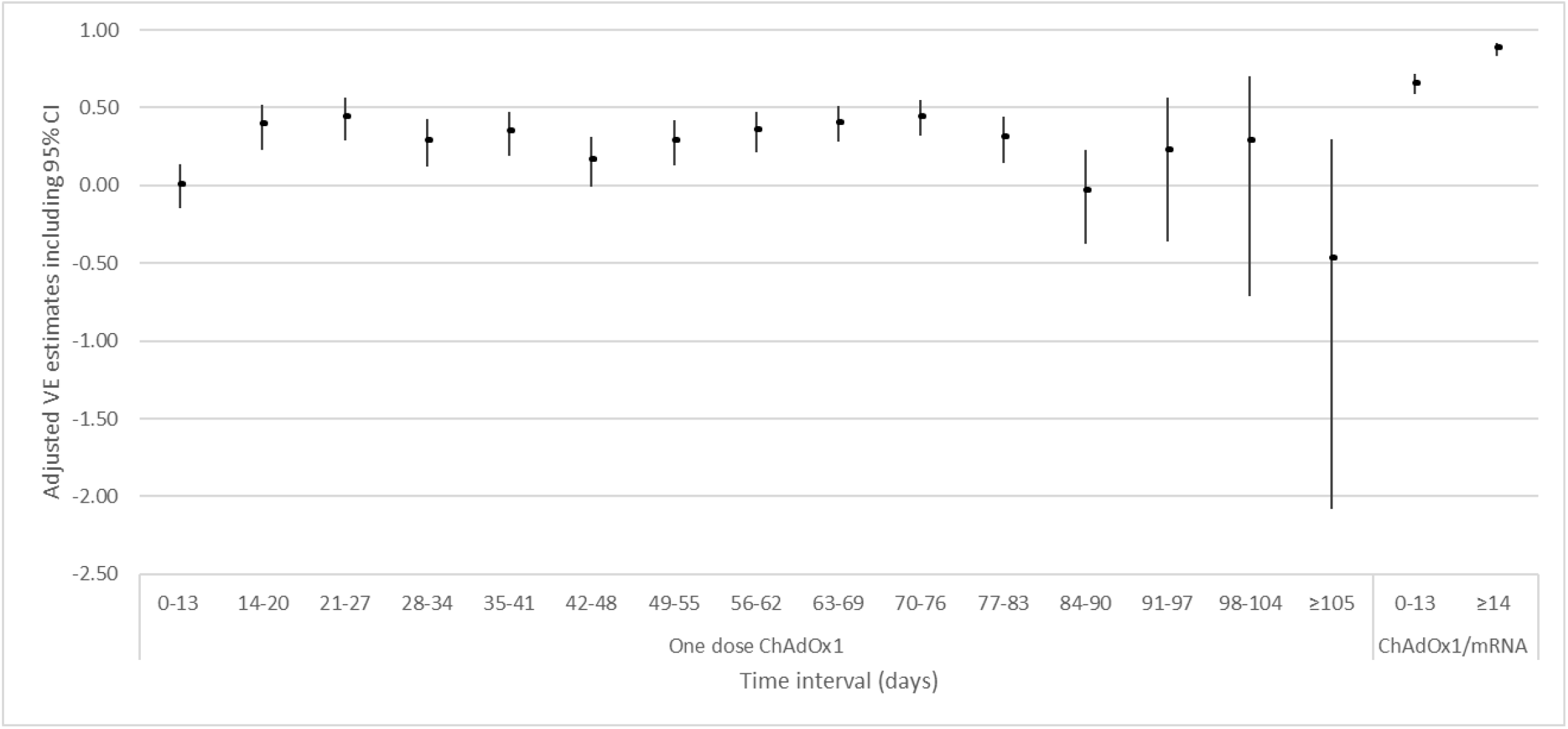
Unadjusted and adjusted VE estimates against RT-PCR SARS-CoV-2 infection of one dose of the ChAdOx1 vaccine and the ChAdOx1/mRNA vaccine schedule.

For all-cause hospitalization, significant adjusted VE estimates were observed at both 0-13 days (VE=36%, 95% CI: 28; 42) and from 14 days (VE=36%, 95% CI: 33; 40) after the first dose of ChAdOx1 and until receiving a second dose of an mRNA vaccine, respectively. Adjusted VE estimates of 43% (95% CI: 36; 49) and 50% (95% CI: 45; 55) was observed at 0-13 days and from 14 days onwards after the ChAdOx1/mRNA vaccine schedule, receptively. A significant adjusted VE of 93% (95% CI: 80; 98) against COVID-19 related hospitalization was observed from 14 days after the first dose and until receiving a second dose of an mRNA vaccine. No COVID-19 related hospitalizations occurred after the ChAdOx1/mRNA vaccine schedule. Therefore, it was not possible to estimate VE estimates against COVID-19 related hospitalization for the ChAdOx1/mRNA vaccine schedule (Table 3).

**Table 3.** Unadjusted and adjusted VE estimates against all-cause and COVID-19 related hospitalization of one dose of the ChAdOx1 vaccine and the ChAdOx1/mRNA vaccine schedule.

|  |  | All-cause hospitalization |  |  |  |  |  |  | COVID-19 related hospitalization |  |  |  |  |  |  |
| --- | --- | --- | --- | --- | --- | --- | --- | --- | --- | --- | --- | --- | --- | --- | --- |
|  |  |  |  |  | Unadjusted |  | Adjusted* |  |  |  |  | Unadjusted |  | Adjusted* |  |
| Vaccine type | Time interval (days) | No. of events | Person-years | Incidence rate | VE, % | 95% CI | VE, % | 95% CI | No. of events | Person-years | Incidence rate | VE, % | 95% CI | VE, % | 95% CI |
| Unvaccinated |  | 149,710 | 1,646,552 | 0.0909 | Reference |  | Reference |  | 1,821 | 1,646,552 | 0.0011 | Reference |  | Reference |  |
| One dose of the ChAdOx1 vaccine | 0-13 | 358 | 5,529 | 0.0648 | 29 | 3; 48 | 36 | 28; 42 | 6 | 5,529 | 0.0011 | 19 | -3,020; 97 | 6 | -110; 58 |
|  | ≥14 | 1,750 | 27,195 | 0.0644 | 29 | 19; 38 | 36 | 33; 40 | 3 | 27,195 | 0.0001 | 90 | -1,220; 100 | 93 | 80; 98 |
| The ChAdOx1/mRNA vaccine schedule | 0-13 | 333 | 5,197 | 0.0641 | 30 | 3; 49 | 43 | 36; 49 | 0 | 5,197 | 0 | - | - | - | - |
|  | ≥14 | 461 | 7,772 | 0.0593 | 35 | 15; 50 | 50 | 45; 55 | 0 | 7,772 | 0 | - | - | - | - |

\*Adjusted for age, sex, heritage and comorbidity

For all-cause death, a significant adjusted VE of 48% (95% CI: 23; 65) was observed at 14 days after one dose of the ChAdOx1 vaccine and until receiving a second dose of an mRNA vaccine. The VE estimates increased to 93% (95% CI: 53; 99) at 0-13 days and 92% (95% CI: 69; 98) at 14 days onwards after an mRNA vaccine as the second dose. No COVID-19 related deaths occurred after receiving a COVID-19 vaccination and therefore a VE could not be estimated (Table 4).

**Table 4.** Unadjusted and adjusted VE estimates against all-cause and COVID-19 related death of one dose of the ChAdOx1 vaccine and the ChAdOx1/mRNA vaccine schedule.

|  |  | All-cause death |  |  |  |  |  |  | COVID-19 related death |  |  |  |  |  |  |
| --- | --- | --- | --- | --- | --- | --- | --- | --- | --- | --- | --- | --- | --- | --- | --- |
|  |  |  |  |  | Unadjusted |  | Adjusted* |  |  |  |  | Unadjusted |  | Adjusted* |  |
| Vaccine type | Time interval (days) | No. of events | Person-years | Incidence rate | VE, % | 95% CI | VE, % | 95% CI | No. of events | Person-years | Incidence rate | VE, % | 95% CI | VE, % | 95% CI |
| Unvaccinated |  | 7,662 | 1,652,106 | 0.0046 | Reference |  | Reference |  | 122 | 1,652,106 | 7.4E-05 | Reference |  | Reference |  |
| One dose of the ChAdOx1 vaccine | 0-13 | 3 | 5,532 | 0.0005 | 88 | -1,030; 100 | 58 | -32; 86 | 0 | 5,532 |  | - | - | - | - |
|  | ≥14 | 26 | 27,260 | 0.0010 | 79 | -3; 96 | 48 | 23; 65 | 0 | 27,260 |  | - | - | - | - |
| The ChAdOx1/mRNA vaccine schedule | 0-13 | 1 | 5,201 | 0.0002 | 96 | -11,190; 100 | 93 | 53; 99 | 0 | 5,201 |  | - | - | - | - |
|  | ≥14 | 2 | 7,782 | 0.0003 | 94 | -1,390; 100 | 92 | 69; 98 | 0 | 7,782 |  | - | - | - | - |

\*Adjusted for age, sex, heritage, comorbidity and hospital admission

## Discussion

This nationwide population-based cohort study showed a significant reduction in the risk of SARS-CoV-2 infection 14 days after the second dose with a VE of 88% (95% CI: 83; 92) when combining the viral vector vaccine ChAdOx1 and an mRNA vaccine. The VE of the ChAdOx1/mRNA vaccine schedule are similar to the VE estimates of 80% (95% CI: 77; 83) and 90% (95% CI: 82; 95) reported in two Danish studies of front-line personnel (healthcare workers) who received two doses of the BNT162b2 mRNA vaccine (16, 17). No COVID-19 related hospitalizations or deaths occurred among individuals who received the ChAdOx1/mRNA vaccine schedule during the study period. This is in accordance with a Danish homologous BNT162b2 mRNA vaccine study (17). This indicates that the combination of the ChAdOx1 and an mRNA vaccine protect against severe outcomes. However, studies with longer follow-up time are needed to confirm these findings, especially because death is a rare outcome in the population of working-age. Immunological studies also indicate that the ChAdOx1/BNT162b2 vaccine schedule is associated with a stronger humoral immune responses and stronger anti-SARS-CoV-2 spike T cell responses compared with two doses of the ChAdOx1 vaccine (18). Also, a 14-day robust humoral and cellular immune response after the second dose of BNT162b2 was observed in individuals primed with ChAdOx1 8-12 weeks earlier (19). Heterologous vaccination schedules studies are important, as several countries want to implement a combined vaccination program with the ChAdOx1 vaccine and an mRNA vaccine to avoid restarting the vaccination schedule with two mRNA vaccines. This is due to both changes in recommendations regarding the use of the ChAdOx1 and to avoid vaccine shortage. Additionally, some countries may need to combine viral vector vaccines with mRNA vaccines to boost immunity.

Due to the decision to withdraw the ChAdOx1 vaccine from the Danish vaccination program, the second vaccine dose was postponed, and it was therefore possible to evaluate the VE of one dose of the ChAdOx1 vaccine. No effectiveness against SARS-CoV-2 infection and COVID-19 related hospital admission during the first 0-13 days after one dose of the ChAdOx1 vaccine was shown. This was expected due to the run-in period before immunity is anticipated to occur. Previous studies have shown similar results of one dose of the ChAdOx1 vaccine (5-7). A test-negative case-control study from England including adults aged 70 years and older showed a VE of 22% (95% CI: 11; 32) from 14-20 days, reaching a VE of 73% (95% CI: 27; 90) at 35 days and onwards after one dose of the ChAdOx1 vaccine (5). Additional protection against hospital admission was observed, suggesting a VE against emergency hospital admission of 80% (5). A cohort study from England, including long-term care facility residents aged 65 years and older, showed adjusted HR against infection immediately after the first dose translated to VE estimates of 49% (95% CI: 01; 74) at 0-6 days, 42% (95% CI: 04; 65) at 7-13 days, 67% (95% CI: 32; 84) at 28-34 days and 68% (95% CI: 34; 85) at 35-48 days (6). A national prospective cohort study from Scotland reported VE estimates against hospital admissions of 70% (95% CI: 63; 76) at 7-13 days, 74% (95% CI: 66; 81) at 14-20 days, 84% (95% CI: 72; 90) at 21-27 days and 94% (95% CI: 73; 99) at 28-34 days after the first dose of the ChAdOx1 vaccine (7). The differences in the VE estimates between these studies (5, 6) and the present study may be explained by differences in the study populations. As of 30^th^ June 2021, Denmark has the highest testing rate for SARS-CoV-2 per 100,000 individuals among European countries (20). A result of the high testing rate we may have detected more SARS-CoV-2 infections in Denmark than in England and Scotland and therefore observe a lower VE of one dose of the ChAdOx1 vaccine. Furthermore, differences in SARS-CoV-2 variants and the background risk of COVID-19 during the study period may also affect the VE estimates. Our results indicate that the ChAdOx1/mRNA vaccine schedule are effective against the alpha variant, which became the dominating variant during the study period.

### Strengths and limitations

The strengths of this study are the high-quality registers and the possibility to use the unique personal identifier to link data on all residents in Denmark. The national testing strategy during the study period, including unlimited access to free-of-charge RT-PCR tests nationwide, led to a high proportion of the population being tested, which enabled us to capture data on both asymptomatic and symptomatic infections. Another strength was the access to national data on all laboratory confirmed RT-PCR SARS-CoV-2 infections. Also, a high sensitivity (97.1%) and specificity (99.98%) was observed for the RT-PCR test (21) ensuring a low risk of misclassification. An effort was made to ensure that all individuals had equal opportunities to receive the COVID-19 vaccines. This was done through an online booking system, special campaigns, offering vaccination in some workplaces, translating the material to other languages than Danish and English and arranging transport for those who were not able to reach the vaccination clinics on their own.

This study has also several limitations. Since we were not able to discriminate between asymptomatic and symptomatic infections, it was not possible to assess the severity of a COVID-19 infection after vaccination based on symptoms. We used a positive SARS-CoV-2 test prior to hospitalization and death (i.e. COVID-19 related hospitalization and death) as proxy for the severity of COVID-19, although this definition might be subject to uncertainty since COVID-19 may not be the cause for these outcomes. Another limitation is that differences in test behavior related to vaccination status may exist, this could result in capturing less asymptomatic infections in vaccinated individuals and lead to elevated VE estimates. The Cox regression models were adjusted for potential confounders including calendar time, age, sex, heritage, hospital admission and comorbidity. However, we cannot eliminate differences in health-seeking behavior or test activity and residual confounding from comorbidity being classified as a dichotomous covariate. This classification does not allow for the exclusion of differences in individual comorbidities across vaccination status, such as the vaccinated cohort being more or less severely ill than the unvaccinated cohort. It was mainly front-line personnel who received the ChAdOx1 vaccine (99.3%). Therefore, we cannot eliminate confounding by indication, assuming that front-line personnel are more exposed to SARS-CoV-2 than the general population (23, 24), which could result in a lower VE in this group. However, front-line personnel are also better trained to use personal protective equipment than the general population, which would moderate the increased risk from their occupational setting. Through the study period, many individuals received COVID-19 vaccines other than ChAdOx1/mRNA vaccines, thus increasing the overall vaccine coverage in the general population, and thereby creating indirect protective herd immunity.

In conclusion, VE against SARS-CoV-2 infection was 88% of the ChAdOx1/mRNA vaccine schedule which is similar to the VE of two doses of the BNT162b2 mRNA vaccine. A single dose of the ChAdOx1 vaccine seems to be protective against SARS-CoV-2 infection after 14 days and up to 83 days, but the second dose of an mRNA vaccine is needed to maintain a significant VE. No COVID-19 related hospitalizations were observed after the second dose, and no COVID-19 related deaths were observed after neither the first dose ChAdOx1 nor the ChAdOx1/mRNA vaccine schedule.

## Supporting information

Table S1. Overview of ICD-10 codes included in the comorbidity variable

Table S2. Definition of heritage

## Data Availability

Because of data protection regulation, data cannot be shared directly by the authors. Data is accessible to researchers from an authorised institution after permission from the Danish Data Protection Agency and Danish Health and Medicines Authority

## Funding

This research did not receive any funding.

## Declaration of interest

All authors declared no competing interest.

## Acknowledgement

The authors are grateful to the Danish Health Data Authority for their help in defining the population. We would also like to thank the Department of Data Integration and Analysis at Statens Serum Institut for data management.

## Notes

### Competing Interest Statement

The authors have declared no competing interest.

### Author Declarations

The data included in this research is part of the Danish national COVID-19 surveillance system database at Statens Serum Institut. The data are available for research upon reasonable request and with permission from the Danish Data Protection Agency and Danish Health and Medicines Authority. Ethics: We used only administrative register data for the study. According to Danish law, ethics approval is exempt for such research, and the Danish Data Protection Agency, which is a dedicated ethics and legal oversight body, thus waives ethical approval for our study of administrative register data, when no individual contact of participants is neccessary and only aggregate results are included as findings. The study is therefore fully compliant with all legal and ethical requirements and there are no further processes available regarding such studies.

