## Supplementary material for "Vaccine effectiveness when combining the ChAdOx1 vaccine as the first dose with an mRNA COVID-19 vaccine as the second dose": Table S1. Overview of ICD-10 codes included in the comorbidity variable

| **Disease** | **ICD-10 codes** |
| --- | --- |
| Diabetes | E10-E14 |
| Adiposity | E65-E68 |
| Cancer | C00-C80, Z85 |
| Neurological diseases | G10-G14, G20-G23, G35-G37, G71-G73, G80-G83, G90-G91, G93-G96, G99, M51 (without G360, G902) |
| Kidney diseases | N18-N200Y, Z992 |
| Hematological cancers | C81-C96, Z856-Z857 |
| Cardiovascular diseases | I20-I21, I230-I259Z, I45-I499Z, I24, I50-I099Z, I340-I399Z, I05, I44-I45, I21-I029Z, I26-I289Z, I40-I439Z, I50-I28Z, R01-R012B, I10-I59Z |
| Respiratory diseases | DJ400-DJ998Z, DJ40, DJ100-DJ229Z, DJ68, DJ430-DJ499 |
| Immune diseases | B200-B249Z, B20, Z21, D800-D899Z, Z923, Z926, Z941-Z949, Z94 (without Z945, Z947) |
| Other diseases | K700-K709, DK70, E150-E909Z, D500-D649Z, D709-D779Z, D50, D650-D699Z, K710-K778Z, Q200-Q349Z, A150-A199Z, A15, Z902, Z905 |
