## Supplementary material for "Vaccine effectiveness when combining the ChAdOx1 vaccine as the first dose with an mRNA COVID-19 vaccine as the second dose": Table S2. Definition of heritage

| **Heritage** | **Definition** |
| --- | --- |
| Danish | Individuals who were born in Denmark or abroad and have at least one parent who is Danish citizen and born in Denmark. |
| Western | Individuals who is affiliated to either Nordic countries, EU countries, Andorra, Liechtenstein, Monaco, San Marino, Switzerland, the United Kingdom, the Vatican City, Canada, the United States, Australia and New Zealand. |
| Non-western | Individuals affiliated to all other countries than the countries defined by western heritage. |
